# Occupational Health and Safety among Healthcare Professionals at the Ashanti Regional Hospital, Ghana

**DOI:** 10.1101/2025.06.05.25329091

**Authors:** Francis Opoku, Jonathan Boakye-Yiadom, Francis Oppong, Thomas Peprah Agyekum, Ebenezer Dassah

## Abstract

**Background:** In Ghana, occupational health and safety (OHS) is vital in protecting healthcare professionals from workplace hazards such as infectious diseases, physical injuries, and psychological stress. These risks can negatively affect staff well-being and productivity. Strengthening OHS practices is essential not only for creating safer work environments but also for improving the quality of healthcare delivery. Despite its importance, OHS implementation in healthcare facilities remains a challenge.

**Objective:** This study assesses the OHS conditions at Ashanti Regional Hospital, Ghana. It identifies common hazards, evaluates healthcare workers’ knowledge, attitudes, and practices (KAP), and analyzes factors influencing these outcomes.

**Methods:** A cross-sectional study was conducted among healthcare professionals at the Ashanti Regional Hospital. Data were collected from 181 participants using a structured questionnaire. The Organizational Network Analysis mobile software (https://ona.io/login) was used for data collection, and statistical analysis was performed using Stata 17.0. Descriptive statistics were summarized as frequencies, percentages, and means. The Mann-Whitney U and Kruskal-Wallis tests were used to compare median KAP scores across groups at a 0.05 significance level with 95% confidence.

**Results:** The average age of participants was 37±6.0 years, with most being female and married. Over half held a diploma qualification. Median KAP scores were significantly higher among professionals who had attended safety training programs (p = 0.026, 0.033, and 0.050, respectively). Additionally, job position significantly influenced knowledge and attitude scores (p = 0.040 and 0.045).

**Conclusion:** The study highlights the importance of targeted safety training programs and department-specific interventions to enhance safety culture in healthcare settings. Healthcare workers who receive proper OHS training demonstrate lower exposure to occupational hazards, emphasizing the need for continuous and updated training. The study recommends improving safety training programs, reinforcing adherence to protective measures, and providing mental health support in the workplace to reduce risks and improve well-being of healthcare professionals.

## 1. Introduction

Occupational Health and Safety (OHS) is an interdisciplinary field of study that aims to protect workers from factors that could compromise their health while they are at work, promote and maintain the highest level of physical, mental, and social well-being for workers in all occupations, and place workers in a workplace that is compatible with their physiological and psychological capabilities [1]. A hazard is a source or situation with a potential for harm in terms of human injury or ill health, damage to property, damage to the environment, or a combination of these. Hazards can include physical, chemical, biological, psychological, and ergonomic factors [2]. Risk is defined as the probability of a specific adverse health effect occurring within a specified period or as a result of a particular exposure situation. It is a function of the probability of an adverse health effect and the severity of that effect, consequential to a hazard(s) in a defined population [2].

Assessment of occupational hazards among healthcare professionals is a critical area of research that aims to identify and mitigate the risks and challenges faced by workers in the healthcare industry. Healthcare professionals, including doctors, and nurses are exposed to a wide range of occupational hazards due to the nature of their work. These hazards can have detrimental effects on the health and well-being of healthcare workers, leading to injuries, illnesses, and even long-term disabilities [3]. Occupational health and safety are important aspects of every workplace, and the healthcare sector is no exception. These risks can include exposure to infectious diseases, physical injuries from heavy lifting or patient handling, chemical and biological hazards, and psychological stress [4].

Every year an estimation of over 2.3 million workers lose their lives through occupational accidents and work-related diseases which means 0.27% of deaths recorded daily globally including healthcare professionals according to the International Labour Organization. Healthcare professionals encompass individuals who engage themselves in actions with their primary intent to enhance health care. Globally more than 50 million individuals are engaged as healthcare professionals [1].

Studies have shown that healthcare professionals have higher rates of work-related injuries and illnesses compared to workers in other industries [5,6]. The physical and psychological toll of these hazards not only affects the well-being of healthcare professionals but can also have an impact on patient care and overall healthcare service delivery [7].

Given the high rates of work-related injuries and illnesses in the healthcare sector, adequate training is essential to equip healthcare professionals with the knowledge and skills needed to identify, prevent, and mitigate occupational hazards. Training programs play a crucial role in raising awareness about potential risks, promoting safe work practices, and empowering healthcare workers to protect themselves and their patients [8].

Researching in this area typically focuses on identifying the various occupational hazards that healthcare professionals are exposed to, and the necessary training needed, evaluating the associated risks, and developing strategies to prevent or minimize them. Common hazards faced by healthcare workers include exposure to infectious diseases, needlestick injuries, musculoskeletal injuries from lifting and transporting patients, exposure to hazardous chemicals and drugs, as well as workplace violence and stress [9]. To ensure the safety and well-being of healthcare professionals and ultimately improve patient care, it is crucial to assess their occupational health and safety training needs. Assessment of these needs involves identifying the knowledge gaps and skill deficiencies that exist in the current training programs available to healthcare professionals [10].

In recent years, the importance of occupational hazard and safety training in the healthcare sector has become even more evident. The COVID-19 pandemic, for example, has highlighted the importance of infection prevention and control practices, personal protective equipment (PPE) usage, and mental health support for healthcare professionals [11]. This has prompted a renewed focus on the assessment of training needs to ensure healthcare professionals are adequately prepared to face such challenges.

## 2. Methods

### 2.1 Study Design

A cross-sectional study design using quantitative methods to assess the occupational health and safety hazards among healthcare professionals within the Ashanti Regional Hospital, Kumasi was adopted. Within four weeks, questionnaires were administered among healthcare professionals. Both exposure variables and the outcome variable were measured at the same point in time.

### 2.2 Study site

Ghana is centrally positioned along the coast of West Africa, covering a total land area of 238,537 square kilometers. This study was conducted in the Ashanti Regional Hospital formally known as Agogo Hospital, which is located in the central part of the country and is the second most populous region after the Greater Accra Region. The region’s healthcare infrastructure consists of 1,654 health facilities, including 1,120 CHPS compounds, 29 clinics, 71 maternity homes, 165 health centres, 5 polyclinics, 26 district hospitals, 125 other hospitals, as well as a regional hospital, a university hospital, and a teaching hospital.

The study was conducted at the Kumasi South Hospital (KSH), which was established in 1976, and originally served as an urban health centre before being designated as the second largest hospital in the southern part of the Ashanti region of Ghana. In 2002, it attained the status of Ashanti Regional Hospital. Catering to a population estimated at 73,864, representing 5.6% of residents of Kumasi Metropolitan and its surroundings, the hospital functions as both a primary care provider and a referral centre. Its jurisdiction extends to neighbouring districts such as Bosomtwe District and Ejisu-Juaben District, where it serves as the principal public healthcare facility. Offering a wide range of medical services, including outpatient care, inpatient treatment, laboratory testing, pharmacy services, maternity care, and various specialized clinics, the hospital is equipped to handle diverse healthcare needs.

Surgical procedures performed at KSH encompass routine operations such as cesarean sections and hernia repairs, as well as specialized interventions including prostatectomy and myomectomy. The general occupational Health & Safety legal and regulatory environment includes fragmented local and international laws and conventions to protect healthcare workers under Ghana Health Service and Teaching Hospitals, 1996 (Act 526).

### 2.3 Study population

All healthcare workers (clinical and non-clinical staff) working at the Kumasi South Hospital constituted the study population. A convenient sampling method, as per the following study inclusion and exclusion criteria, was used to select study participants from the various directorate/unit levels. To ensure accurate and relevant data collection on occupational health and safety conditions, only those currently working at the hospital during the study period were eligible to participate. Healthcare professionals on leave at the time of data collection were excluded to maintain the integrity of real-time workplace hazard assessments. Additionally, individuals who declined to provide consent were not included, in alignment with ethical considerations and the principles of voluntary participation.

### 2.4 Study instrument

The structure questionnaire consists of four main parts: Socio-Demographic Information: This section includes nine items assessing participants’ background characteristics: age, sex, marital status, educational background, job position, work schedule, years of work experience, unit/department of work, and the need for additional occupational health and safety (OHS) training. Knowledge Assessment: Five items evaluate participants’ awareness and understanding of OHS-related aspects, including the availability of OHS guidelines, emergency procedures, proper use of first aid equipment, fire outbreak management, and precautions for handling toxic and hazardous substances. Attitudes Toward OHS: This section consists of ten items assessing participants’ perceptions and attitudes regarding workplace safety, teamwork safety, adherence to safety regulations, the effectiveness of safety training, and the role of personal protective equipment (PPE) in infection prevention. Safety Practices: Five items measure participants’ adherence to safety protocols, including workplace organization, proper use of PPE, handling of contaminated and sharp materials, hand hygiene, and the reporting and management of needle stick injuries. Responses were recorded using a 5-point Likert scale, with options ranging from (1) Never, (2) Rarely, (3) Sometimes, (4) Often, to (5) Always for knowledge and attitudes.

### 2.5 Ethical considerations

The study protocols were approved by the Committee on Human Research Publications and Ethics (CHRPE) at Kwame Nkrumah University of Science and Technology (KNUST), Kumasi, Ghana (Approval Number: CHRPE/AP/1012/24). Written informed consent was obtained from all participants before data collection.

### 2.6 Data Collection Procedure

Data were collected using structured questionnaires through electronic tablets and smartphones, utilizing a specialized software application called Organizational Network Analysis (ONA), designed specifically for Android platforms, from October 1, to November 20, 2024. To streamline data entry into the ONA database, the questionnaire was formatted for electronic input and securely uploaded to a dedicated cloud server, which was encrypted with a password.

This cloud server had a dual purpose: it hosted and stored all collected data, accessible only with a password known exclusively to the principal investigator. Each research assistant was equipped with either an electronic tablet or smartphone, preloaded with a customized version of the ONA software. This setup allowed consistent communication with the cloud server, facilitating easy access to the questionnaires. All respondent data was transmitted to the server and compiled for subsequent analysis.

### 2.7 Data Analysis

Data were analyzed using Stata version 17 and cleaned using Microsoft excel. Descriptive statistics summarized the demographic information and pre-training knowledge levels. For inferential analysis, non-parametric tests were employed to assess associations between occupational hazards and demographic variables. Specifically, the Mann-Whitney U Test was used to compare median scores of knowledge, attitude, and practices between two independent groups, while the one-way Kruskal-Wallis Test was applied for comparisons across more than two groups. These tests were selected due to violations of normality assumptions in the data. All statistical tests were two-tailed and conducted at a 5% significance level with a 95% confidence interval. All statistical tests were conducted at a significance level of 0.05 with a 95% confidence level.

## 3. Results

### 3.1 Sociodemographic characteristics of healthcare professionals

The socio demographics characteristics of 181 healthcare workers, showing that the majority are female (74.30%), married (69.06%), diploma holders (53.59%), and nurses (48.31%), with most working permanent day shifts (54.80%) and having 1-5 years of experience (32.95%); additionally, 68.33% adhere to safety precautions **(Table 1).**

**Table 1:** Sociodemographic characteristics of healthcare workers.

| Variables | Frequency (n=181) | Percentage (%) |
| --- | --- | --- |
| Age (years) | X±SD:37±6.0 |  |
| 37 and below | 100 | 55.25 |
| Above 37 | 81 | 44.75 |
| Sex |  |  |
| Male | 46 | 25.70 |
| Female | 133 | 74.30 |
| Marital Status |  |  |
| Single | 48 | 26.52 |
| Married | 125 | 69.06 |
| Divorce | 8 | 4.42 |
| Highest Level of Education |  |  |
| Certificate | 13 | 7.18 |
| Diploma | 97 | 53.59 |
| First Degree | 56 | 30.94 |
| Second Degree | 11 | 6.08 |
| Fellowship | 4 | 2.21 |
| Job Position |  |  |
| Doctor | 27 | 15.17 |
| Nurse | 86 | 48.31 |
| Midwife | 48 | 26.97 |
| Medical Technicians | 9 | 5.06 |
| Public Health Officer | 8 | 4.49 |
| Work Schedule |  |  |
| Alternate between day and night shift | 77 | 43.50 |
| Permanent day shift only | 97 | 54.80 |
| Permanent night shift only | 3 | 1.69 |
| Working Experience (years) |  |  |
| 1-5 | 58 | 32.95 |
| 6-10 | 56 | 31.82 |
| 11-15 | 32 | 18.18 |
| 16-20 | 21 | 11.93 |
| >20 | 9 | 5.11 |
| Unit/department of Work |  |  |
| Out patient's department (OPD) | 32 | 17.88 |
| Oral Health | 7 | 3.91 |
| Emergency | 27 | 15.08 |
| Obstetrics and Gynecology (O&G) | 21 | 11.73 |
| Eye | 6 | 3.35 |
| Surgical | 3 | 1.68 |
| Medical Laboratory | 42 | 23.46 |
| Child health | 26 | 14.53 |
| Mental health | 13 | 7.26 |
| Public health | 2 | 1.12 |
| Attendance of safety precautions meetings |  |  |
| Yes | 123 | 68.33 |
| No | 57 | 31.67 |

**Source: Author's Construct, 2024**

### 3.2 Types of occupational hazards faced by healthcare professionals

Figure 1 illustrates the percentage of healthcare professionals experiencing different occupational hazards. The majority (91.11%) faced biological hazards, followed by physical hazards (66.85%), chemical hazards (55.87%), and psychological hazards (41.11%). This indicates that biological hazards are the most prevalent risk in healthcare settings, while psychological hazards are the most commonly avoided.

**Figure 1:**
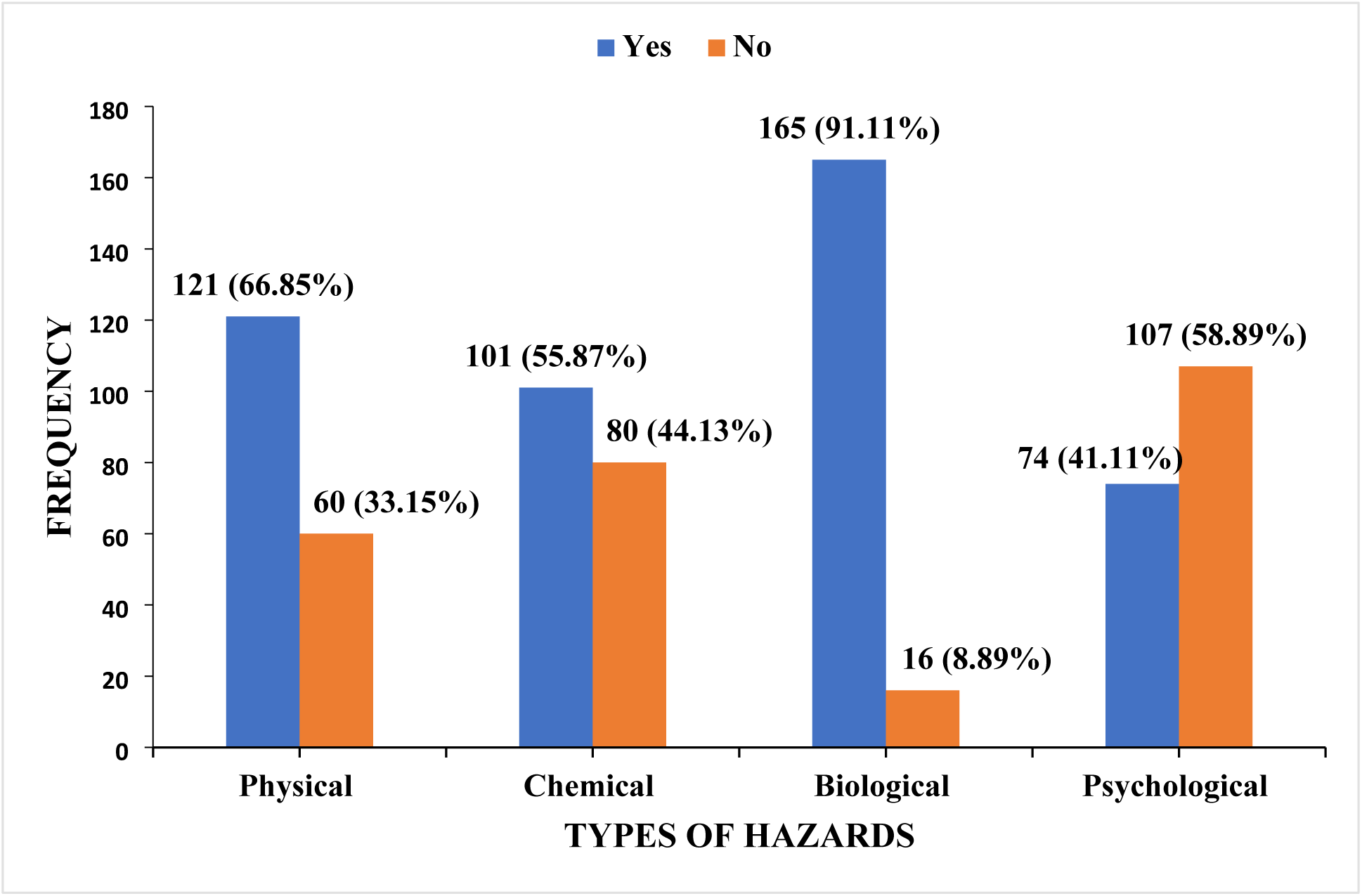
Types of Occupational Hazard faced by healthcare professionals. Source: Author’s Construct, 2024

**Table 2** outlines the risks of healthcare sector workers by risks associated and are categorized as physical, chemical, biological, and psychological risks. The data highlights that healthcare workers face significant occupational hazards, including physical pain such as joint pain 43.80%, chemical exposure (67.62% to medications), biological risks (79.31% experiencing needlestick injuries), and psychological challenges (17.78% reporting burnout), with limited access to mental health support (87.71%).

**Table 2:**
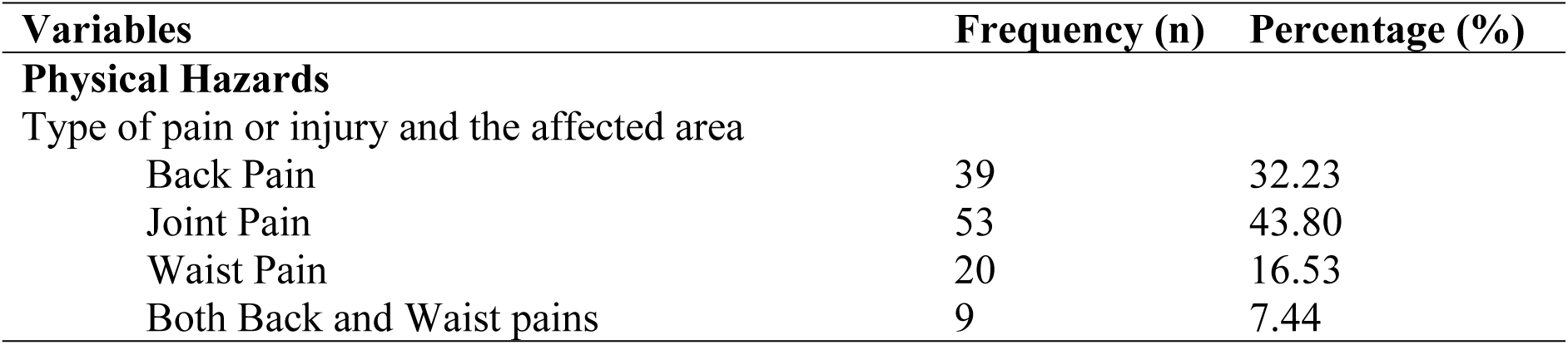
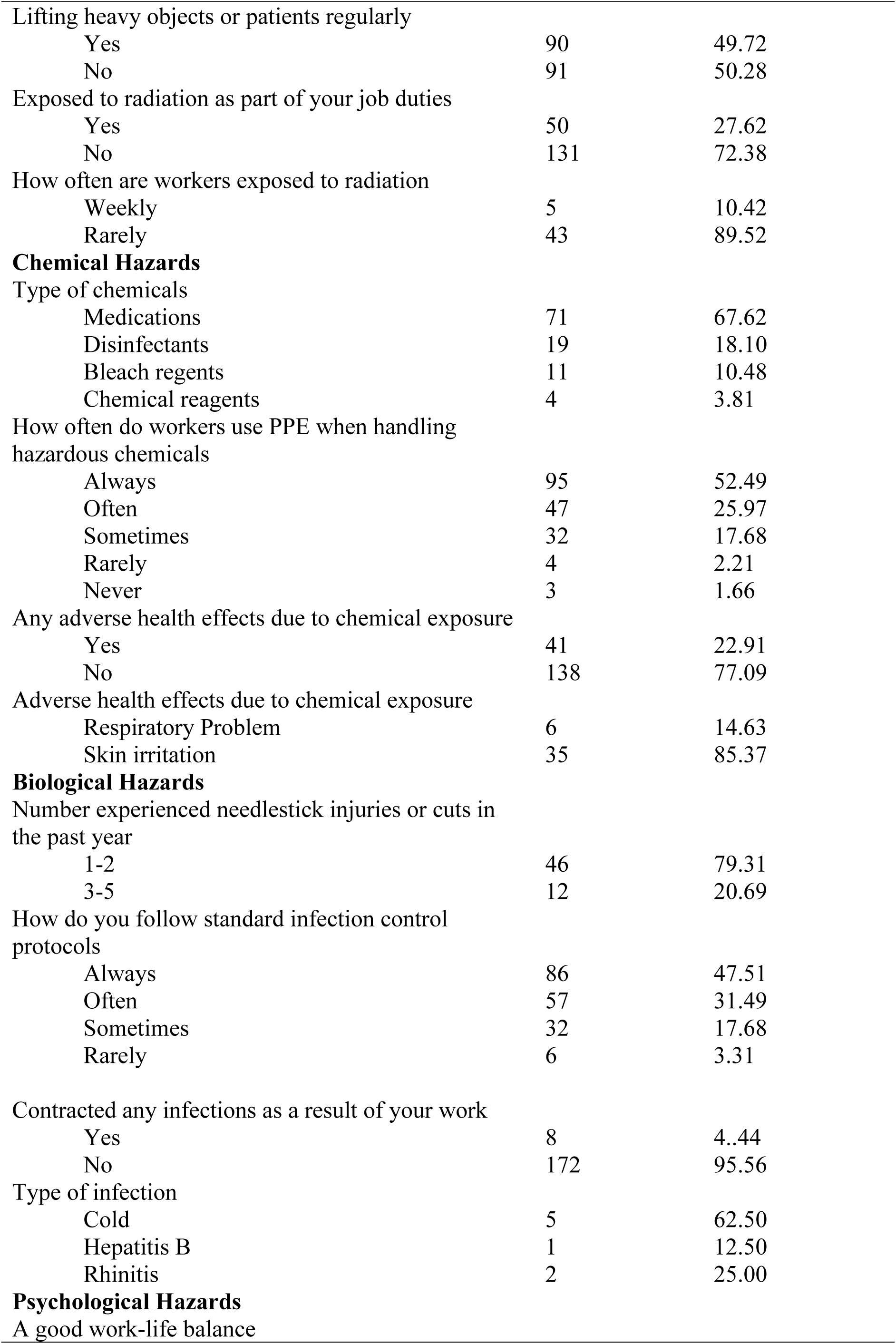
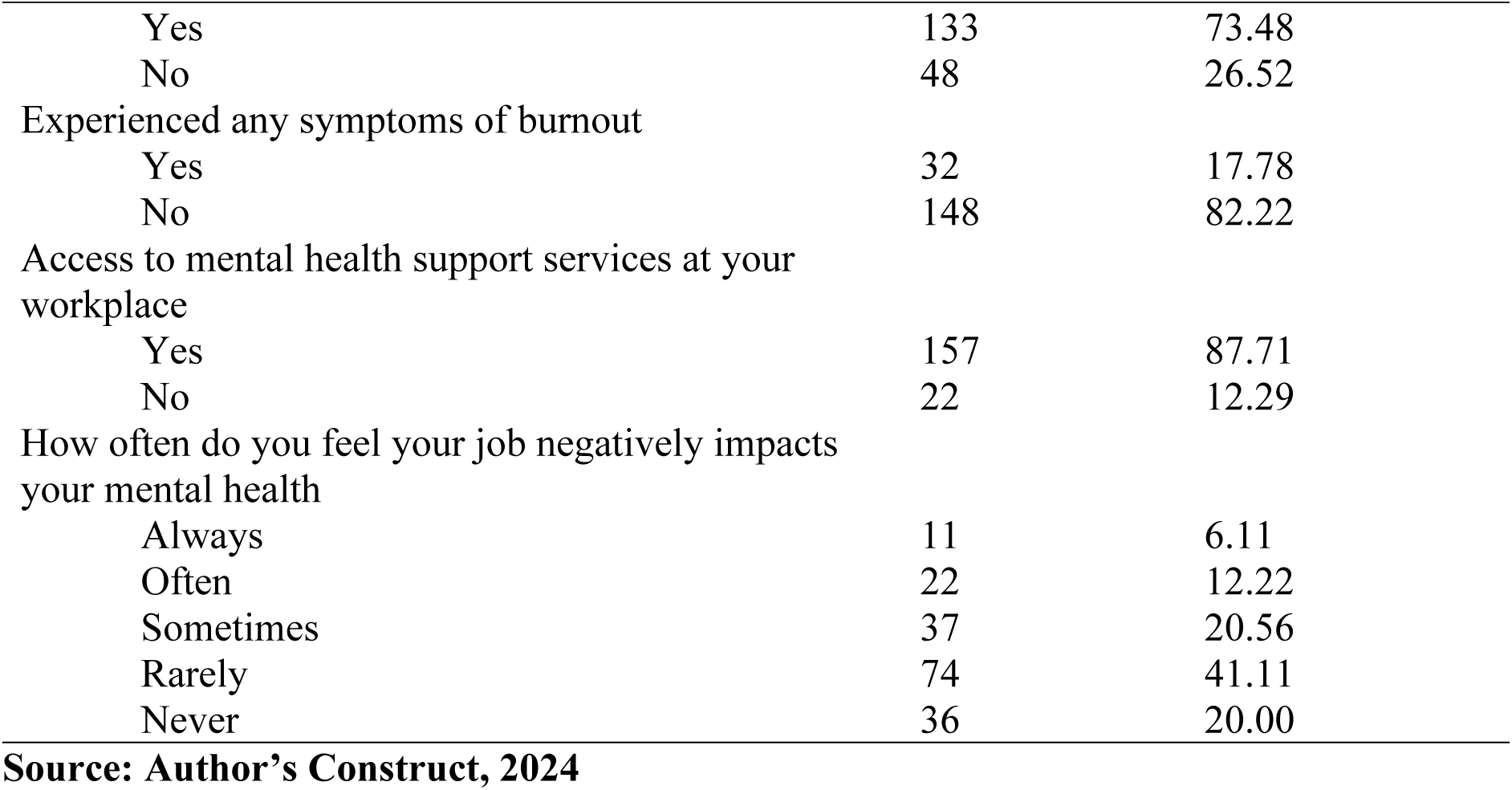
Occupational Hazards Faced by Healthcare Professionals.

| Variables | Frequency (n) | Percentage (%) |
| --- | --- | --- |
| **Physical Hazards** |  |  |
| Type of pain or injury and the affected area |  |  |
| Back Pain | 39 | 32.23 |
| Joint Pain | 53 | 43.80 |
| Waist Pain | 20 | 16.53 |
| Both Back and Waist pains | 9 | 7.44 |

| **Variables** | **Frequency (n)** | **Percentage (%)** |
| --- | --- | --- |
| Lifting heavy objects or patients regularly |  |  |
| Yes | 90 | 49.72 |
| No | 91 | 50.28 |
| Exposed to radiation as part of your job duties |  |  |
| Yes | 50 | 27.62 |
| No | 131 | 72.38 |
| How often are workers exposed to radiation |  |  |
| Weekly | 5 | 10.42 |
| Rarely | 43 | 89.52 |
| **Chemical Hazards** |  |  |
| Type of chemicals |  |  |
| Medications | 71 | 67.62 |
| Disinfectants | 19 | 18.10 |
| Bleach regents | 11 | 10.48 |
| Chemical reagents | 4 | 3.81 |
| How often do workers use PPE when handling hazardous chemicals |  |  |
| Always | 95 | 52.49 |
| Often | 47 | 25.97 |
| Sometimes | 32 | 17.68 |
| Rarely | 4 | 2.21 |
| Never | 3 | 1.66 |
| Any adverse health effects due to chemical exposure |  |  |
| Yes | 41 | 22.91 |
| No | 138 | 77.09 |
| Adverse health effects due to chemical exposure |  |  |
| Respiratory Problem | 6 | 14.63 |
| Skin irritation | 35 | 85.37 |
| **Biological Hazards** |  |  |
| Number experienced needlestick injuries or cuts in the past year |  |  |
| 1-2 | 46 | 79.31 |
| 3-5 | 12 | 20.69 |
| How do you follow standard infection control protocols |  |  |
| Always | 86 | 47.51 |
| Often | 57 | 31.49 |
| Sometimes | 32 | 17.68 |
| Rarely | 6 | 3.31 |
| Contracted any infections as a result of your work |  |  |
| Yes | 8 | 4.44 |
| No | 172 | 95.56 |
| Type of infection |  |  |
| Cold | 5 | 62.50 |
| Hepatitis B | 1 | 12.50 |
| Rhinitis | 2 | 25.00 |
| **Psychological Hazards** |  |  |
| A good work-life balance |  |  |
| Yes | 133 | 73.48 |
| No | 48 | 26.52 |
| Experienced any symptoms of burnout |  |  |
| Yes | 32 | 17.78 |
| No | 148 | 82.22 |
| Access to mental health support services at your workplace |  |  |
| Yes | 157 | 87.71 |
| No | 22 | 12.29 |
| How often do you feel your job negatively impacts your mental health |  |  |
| Always | 11 | 6.11 |
| Often | 22 | 12.22 |
| Sometimes | 37 | 20.56 |
| Rarely | 74 | 41.11 |
| Never | 36 | 20.00 |

**Source: Author's Construct, 2024**

### 3.4 Knowledge of occupational health and safety among healthcare professionals

The majority of healthcare workers demonstrated a high level of awareness regarding occupational health and safety (OHS) procedures, emergency protocols, and hazard management. The proportion of respondents (42.78%) strongly agreed that there are OHS instructions in their working area, with a median response of 4 (IQR: 3, 5), indicating generally positive perceptions. Similarly, 48.07% strongly agreed that emergency procedures are available in their work area, with a median of 4 (IQR: 4, 5), showing high awareness of emergency protocols. An overwhelming majority (72.32%) strongly agreed that they know how to use first aid equipment in emergencies, with a median of 5 (IQR: 4, 5), reflecting strong confidence in first aid knowledge. Likewise, 70.56% strongly agreed that they know what to do in case of chemical contact, and 71.84% strongly agreed they know how to respond to burns or hot object exposure, both with medians of 5 (IQR: 4, 5), indicating a high level of preparedness for specific workplace hazards. This suggest that most workers possess substantial knowledge of safety procedures and emergency responses in their workplace **(Table 3).**

**Table 3:** Knowledge of occupational health and safety among healthcare workers.

| Items | Never<br>n(%) | Rarely<br>n(%) | Sometimes<br>n(%) | Often<br>n(%) | Usually<br>n(%) | N | M<br>(IQR) |
| --- | --- | --- | --- | --- | --- | --- | --- |
| There are instructions for occupational health and safety in your working area | 2(1.11) | 26(14.44) | 41(22.78) | 34(18.89) | 77(42.78) | 180 | 4(3, 5) |
| There are procedures for emergencies inside your working area | 1(0.55) | 11(6.08) | 33(18.23) | 49(27.07) | 87(48.07) | 181 | 4(4, 5) |
| I know how to use first aid equipment in terms of emergency | 0(0.00) | 2(1.13) | 13(7.34) | 34(19.21) | 128(72.32) | 177 | 5(4, 5) |
| I know what to do if I get in contact with a chemical | 0(0.00) | 5(2.78) | 25(13.89) | 23(12.78) | 127(70.56) | 180 | 5(4, 5) |
| I know how to deal when exposed to a burn or contact with a hot object | 1(0.57) | 6(3.45) | 22(12.64) | 20(11.49) | 125(71.84) | 174 | 5(4, 5) |

Data are expressed as percentages and frequencies. M: Median scores, IQR: 25<sup>th</sup> and 75<sup>th</sup> Interquartile266 **Source: Author's Construct, 2024**

### 3.5 Attitude of occupational health and safety among healthcare workers

The data presented in **Table 4** reflect respondents’ perceptions and practices regarding occupational health and safety in a hospital setting. Notably, 69.66% of respondents strongly agreed that they feel safe working with colleagues in a team, and 71.02% strongly agreed that they consider colleagues’ views on hospital-related dangers. A significant proportion also reported benefiting from past experiences to avoid mistakes (60.56%) and acknowledged that clear safety rules reduce accidents (79.21%). Furthermore, 83.80% strongly agreed that safety training is impactful, and 82.12% expressed strong concern for the safety of others. The use of personal protective equipment was affirmed by 73.84% of respondents. Across all items, the median response was consistently 5, with narrow interquartile ranges, indicating a high level of consensus and a strong safety culture among the participants **(Table 4).**

**Table 4:**
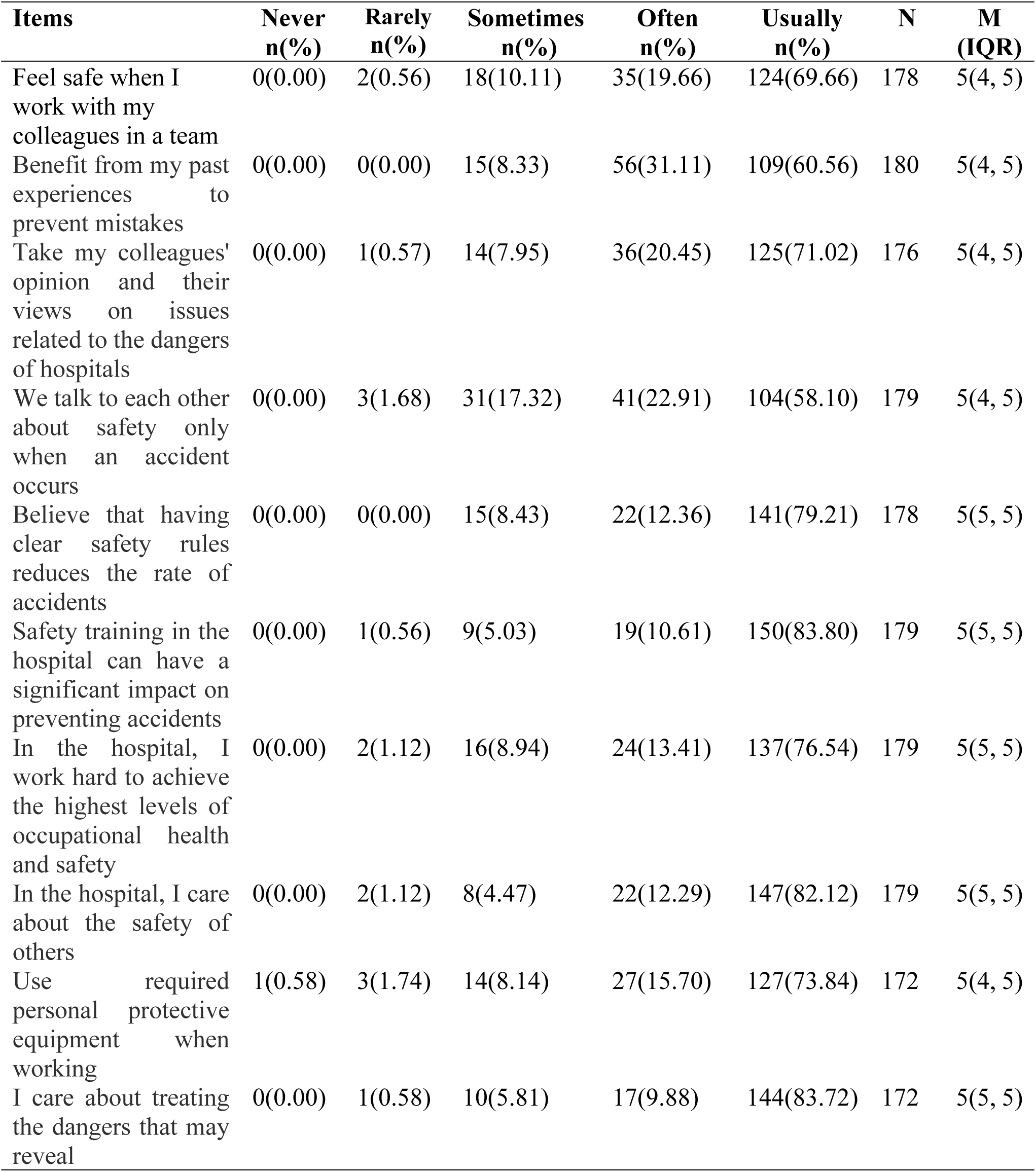
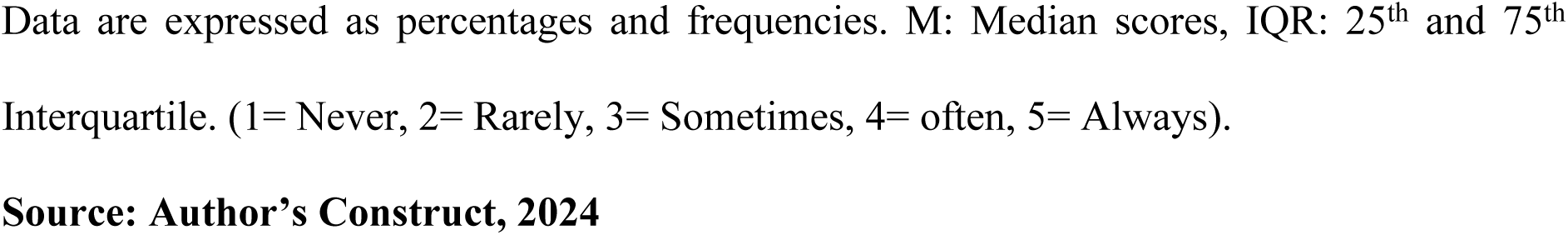
Attitude of occupational health and safety among healthcare workers.

| Items | Never<br>n(%) | Rarely<br>n(%) | Sometimes<br>n(%) | Often<br>n(%) | Usually<br>n(%) | N | M<br>(IQR) |
| --- | --- | --- | --- | --- | --- | --- | --- |
| Feel safe when I work with my colleagues in a team | 0(0.00) | 2(0.56) | 18(10.11) | 35(19.66) | 124(69.66) | 178 | 5(4, 5) |
| Benefit from my past experiences to prevent mistakes | 0(0.00) | 0(0.00) | 15(8.33) | 56(31.11) | 109(60.56) | 180 | 5(4, 5) |
| Take my colleagues' opinion and their views on issues related to the dangers of hospitals | 0(0.00) | 1(0.57) | 14(7.95) | 36(20.45) | 125(71.02) | 176 | 5(4, 5) |
| We talk to each other about safety only when an accident occurs | 0(0.00) | 3(1.68) | 31(17.32) | 41(22.91) | 104(58.10) | 179 | 5(4, 5) |
| Believe that having clear safety rules reduces the rate of accidents | 0(0.00) | 0(0.00) | 15(8.43) | 22(12.36) | 141(79.21) | 178 | 5(5, 5) |
| Safety training in the hospital can have a significant impact on preventing accidents | 0(0.00) | 1(0.56) | 9(5.03) | 19(10.61) | 150(83.80) | 179 | 5(5, 5) |
| In the hospital, I work hard to achieve the highest levels of occupational health and safety | 0(0.00) | 2(1.12) | 16(8.94) | 24(13.41) | 137(76.54) | 179 | 5(5, 5) |
| In the hospital, I care about the safety of others | 0(0.00) | 2(1.12) | 8(4.47) | 22(12.29) | 147(82.12) | 179 | 5(5, 5) |
| Use required personal protective equipment when working | 1(0.58) | 3(1.74) | 14(8.14) | 27(15.70) | 127(73.84) | 172 | 5(4, 5) |
| I care about treating the dangers that may reveal | 0(0.00) | 1(0.58) | 10(5.81) | 17(9.88) | 144(83.72) | 172 | 5(5, 5) |

Data are expressed as percentages and frequencies. M: Median scores, IQR: 25<sup>th</sup> and 75<sup>th</sup> Interquartile. (1= Never, 2= Rarely, 3= Sometimes, 4= often, 5= Always).
**Source: Author's Construct, 2024**

### 3.6 Practices of occupational health and safety among healthcare workers

The table below presents the distribution of responses regarding adherence to occupational health and safety practices among respondents. A significant majority (45.56%) reported always adhering to wearing a medical mask while working, with a median score of 4 and an interquartile range (IQR: 3, 5). Most respondents (83.71%) indicated that they always handle contaminated materials correctly to prevent hazards, resulting in a high median score of 5 and an (IQR 5, 5), indicating strong consistency in responses. Similarly, 60.11% always wore appropriate personal protective equipment (PPE) during working hours, with a median of 4 and IQR of 1. Wearing shoes that cover the foot was also widely practiced, with 62.5% indicating they always did so, yielding a median of 5 (IQR: 4, 5). Finally, adherence to professional rules for disposing of medical waste was the most consistent, with 86.29% of respondents always following these rules, a median of 5, and no variability (IQR: 5, 5), highlighting a strong commitment to proper waste disposal practices among participants **(Table 5).**

**Table 5:**
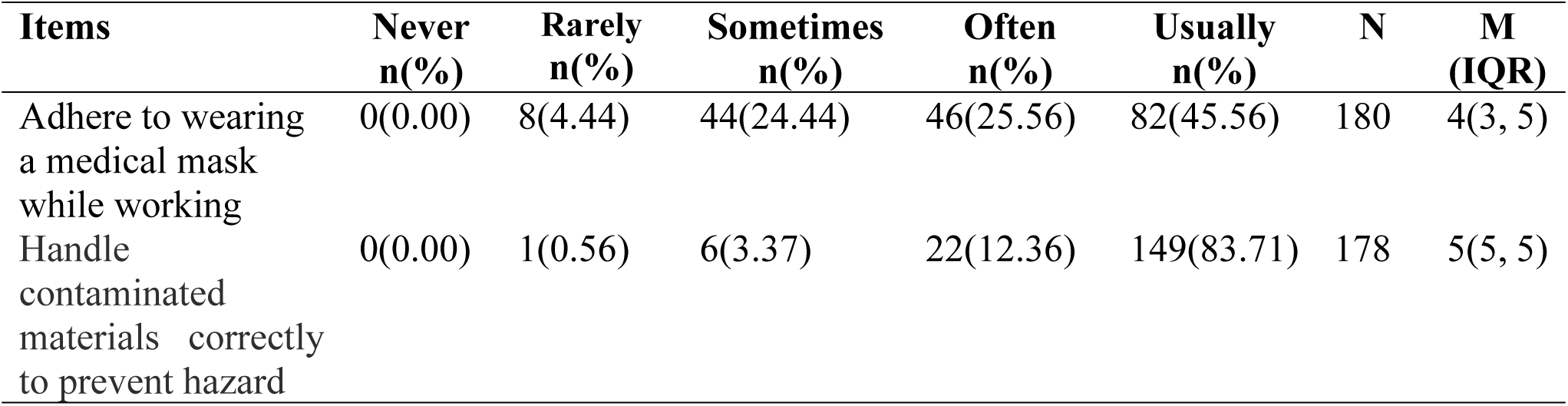
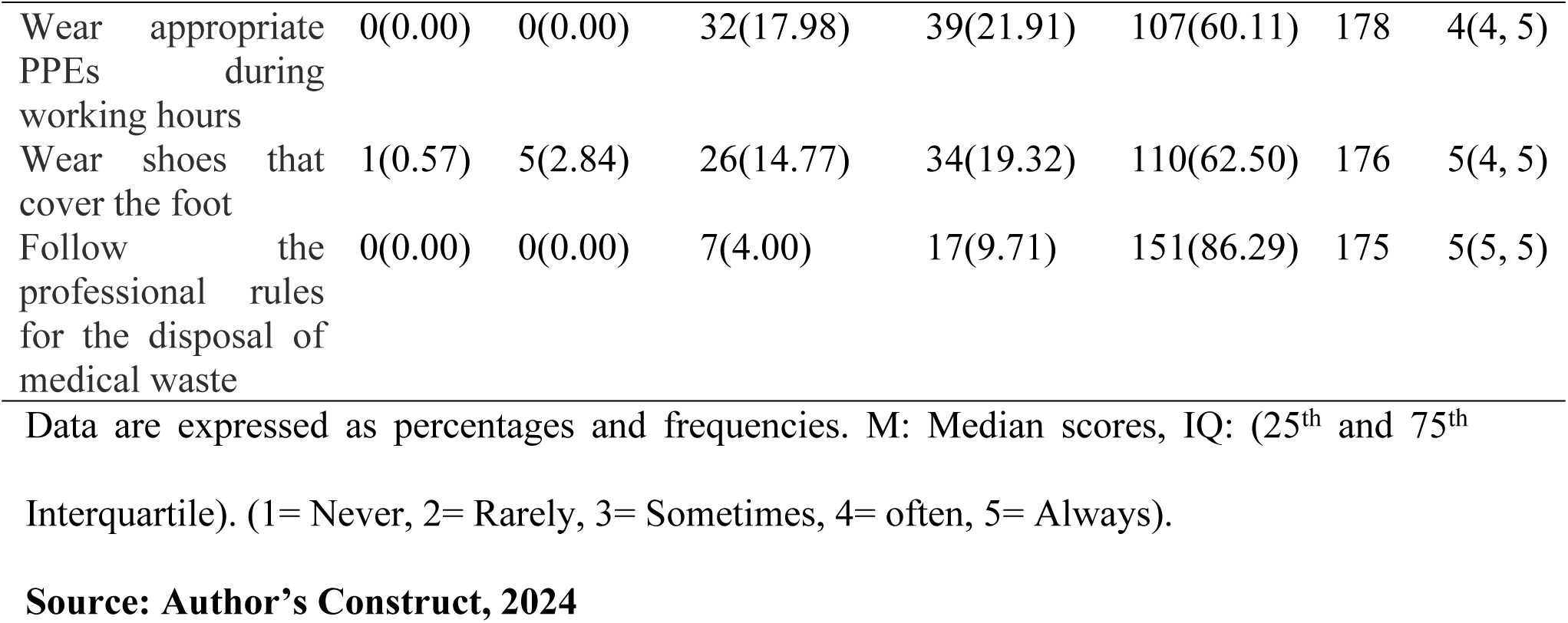
Practices of occupational health and safety among healthcare workers.

### 3.7 Characteristics of the healthcare professionals to knowledge, attitude, and practice of occupational health and safety

The relationship between healthcare professionals’ characteristics and their knowledge, attitude, and practice of occupational health and safety (OHS), highlights significant differences in job position, department, and safety precaution training attendance. Job position had a highly significant effect, both on knowledge (*P* = 0.040) and attitude (p=0.045). Midwives had the highest median knowledge score, 23.5 (IQR, 5), and attitude score 48.0 (IQR, 4) compared to public health officers who scored the lowest median 19.5 (IQR, 6) for knowledge; M=45.5, IQR: 41.5, and 50 for attitude.

A Kruskal-Wallis test was conducted to examine the association between department of work and knowledge levels, revealing a statistically significant relationship (χ² = 23.717, *P* = 0.005). Additionally, the test indicated a near-significant association with attitude levels (χ² = 16.702, *P* = 0.053), suggesting a potential trend worth further investigation.

The highest knowledge scores were observed in the Obstetrics and Gynecology (O&G) unit workers 24.0 (IQR: 3), while that of public health unit workers were lowest 18.5 (IQR: 5) Oral Health 49.0 (IQR: 2) and O&G unit 48.0 (IQR: 4) workers had the highest attitude scores, while public health workers had again the lowest attitude scores 42.0 (IQR: 0). Attendance of safety precaution training significantly impacted all of the three domains knowledge (*P*=0.026), attitude (*P*=0.033) and practice (*P*=0.015).

There was no significant difference in knowledge, attitude, or practice and the other variables like age, sex, marital status, educational level, work schedule, and years of experience. These results underscore the importance of job roles, differences between departments, and training in the development of OHS awareness and practices among health professionals **(Table 6).**

**Table 6:** Characteristics of the healthcare professionals to knowledge, attitude, and practice of occupational health and safety.

| Variables | Knowledge<br>M: (IQR) | P-value | Attitude<br>M: (IQR) | P-value | Practices<br>M: (IQR) | P-value |
| --- | --- | --- | --- | --- | --- | --- |
| Age (years) |  | 0.497 |  | 0.409 |  | 0.692 |
| 37 and below | 22.0 (19, 24) |  | 47.0 (43, 49) |  | 23.0 (20, 25) |  |
| Above 37 | 22.0 (20, 24) |  | 47.0 (42, 49) |  | 23.0 (20, 24) |  |
| Sex |  | 0.376 |  | 0.722 |  | 0.413 |
| Male | 21.5 (19, 24) |  | 47.0 (44, 50) |  | 23.0 (20, 25) |  |
| Female | 22.0 (19, 24) |  | 47.0 (42, 49) |  | 23.0 (20, 25) |  |
| Marital Status |  | 0.066 |  | 0.794 |  | 0.333 |
| Single | 21.0 (19, 23) |  | 47.0 (42.5, 49) |  | 22.0 (19, 24.5) |  |
| Married | 22.0 (19, 24) |  | 47.0 (42, 49) |  | 23.0 (20, 25) |  |
| Divorce | 23.5(22.5, 24.5) |  | 48.0(43.5, 49.5) |  | 23.0 (22, 23.5) |  |
| Highest Level of Education |  | 0.462 |  | 0.058 |  | 0.552 |
| Certificate | 19.0 (19, 22) |  | 42.0 (39, 46) |  | 22.0 (19, 25) |  |
| Diploma | 22.0 (19, 24) |  | 47.0 (45, 49) |  | 23.0 (21, 25) |  |
| First Degree | 21.5 (20, 24) |  | 46.0 (41, 49.5) |  | 22.0 (19, 24) |  |
| Second Degree | 24.0 (19, 24) |  | 47.0 (36, 49) |  | 22.0 (19, 25) |  |
| Fellowship | 23.5 (18, 24.5) |  | 48.0 (41.5, 50) |  | 24.5 (20.5, 25) |  |
| Job Position |  | **0.040\*** |  | **0.045\*** |  | 0.231 |
| Doctor | 23.0 (18, 24) |  | 47.0 (37, 50) |  | 23.0 (19, 25) |  |
| Nurse | 21.0 (19, 23) |  | 46.0 (41, 49) |  | 23.0 (20, 25) |  |
| Midwife | 23.5 (20, 25) |  | 48.0 (46, 50) |  | 23.0 (22, 25) |  |
| Medical Technicians | 22.0 (20, 24) |  | 44.0 (42, 47) |  | 21.0 (20, 24) |  |
| Public Health Officer | 19.5 (17, 23) |  | 45.5 (41.5, 50) |  | 21.0(19.5, 23) |  |
| Work schedule |  | 0.756 |  | 0.581 |  | 0.311 |
| Alternate between day and night shift | 22.0 (19, 24) |  | 47.0 (45, 49) |  | 23.0 (21, 25) |  |
| Permanent day shift only | 21.0 (19, 24) |  | 47.0 (42, 50) |  | 22.0 (20, 25) |  |
| Permanent night shift only | 22.0 (18, 22) |  | 44.0 (39, 48) |  | 23.0 (19,24) |  |
| Working experience (years) |  | 0.464 |  | 0.237 |  | 0.966 |
| 1-5 | 21.0 (19, 24) |  | 47.0 (43, 49) |  | 23.0 (20, 25) |  |
| 6-10 | 22.0 (19, 25) |  | 47.0 (44, 50) |  | 22.5 (20, 25) |  |
| 11-15 | 22.5 (18.5, 24) |  | 46.0 (39, 48) |  | 23.0 (20, 25) |  |
| 16-20 | 21.0 (20, 24) |  | 48.0 (42, 50) |  | 23.0 (20, 24) |  |
| >20 | 23.0 (20, 24) |  | 47.0 (39, 47) |  | 22.0 (22, 24) |  |
| Unit/department of Work |  | **0.005\*** |  | **0.053\*** |  | 0.337 |
| Out patient's department (OPD) | 20.0 (18.5, 23.5) |  | 46.0 (39, 49.5) |  | 22.0 (20, 24) |  |
| Oral Health | 21.0 (21, 23) |  | 49.0 (48, 50) |  | 23.0 (23, 25) |  |
| Emergency | 23.0 (19, 24) |  | 48.0 (42, 49) |  | 23.0 (19, 24) |  |
| Obstetrics and Gynecology (O&G) | 24.0 (22, 25) |  | 48.0 (46, 50) |  | 24.0 (22, 25) |  |
| Eye | 19.5 (16, 24) |  | 45.5 (44, 48) |  | 23.0 (22, 25) |  |
| Surgical | 24.0 (19, 25) |  | 45.0 (45, 47) |  | 22.0 (20, 25) |  |
| Medical Laboratory | 21.0 (19, 23) |  | 46.0 (41, 49) |  | 22.0 (20, 25) |  |
| Child health | 24.0 (21, 25) |  | 48.0 (45, 47) |  | 23.0 (21, 25) |  |
| Mental health | 20.0 (18, 22) |  | 45.0 (40, 47) |  | 23.0 (18, 25) |  |
| Public health | 18.5 (16, 21) |  | 42.0 (42, 42) |  | 17.0 (17, 17) |  |
| Attendance of safety Precautions meeting |  | **0.026\*** |  | **0.033\*** |  | **0.015\*** |
| Yes | 22.0 (19, 24) |  | 47.0 (41, 49) |  | 23.0 (20, 25) |  |
| No | 22.0 (19, 24) |  | 47.0 (44, 49) |  | 22.0 (20, 24) |  |

Data are expressed as M: Median scores, IQR: (25<sup>th</sup> and 75<sup>th</sup> Interquartile) for continuous variables. The differences between medians were tested using the independent sample Mann-Whitney U Test and one-way Kruskal-Wallis Test.
**Source: Author's Construct, 2024**

## 4. Discussion

This study provides a comprehensive analysis of occupational hazards faced by healthcare workers at Kumasi South Hospital, revealing a multifactorial burden involving physical, chemical, biological, and psychological risks, as well as varying levels of knowledge and adherence to occupational health and safety (OHS) practices. The findings point to systemic and contextual factors shaping workplace safety.

One of the prominent findings is the prevalence of physical hazards, especially those resulting from manual patient handling and physical strain during routine clinical activities. This observation aligns with a study at Ho, indicating that healthcare workers are at an elevated risk for musculoskeletal disorders due to the nature of their work [12]. However, a study from the United States, which is more resourced reports lower physical injury rates, attributed to wider adoption of assistive technologies and structured ergonomic training programs [13]. The persistence of these hazards, despite awareness campaigns, may reflect systemic gaps in ergonomic design, insufficient training on proper lifting techniques, and inadequate staffing levels that increase physical workload. Addressing these risks calls for structural interventions such as ergonomic assessments, procurement of lifting equipment, and routine manual handling training, which can collectively reduce strain injuries and promote workforce sustainability.

Chemical hazards appeared as another significant threat, with healthcare workers commonly exposed to disinfectants, medications, and other hazardous substances. This aligns with findings in similar low- and middle-income countries, where chemical exposure has been linked to chronic disease such as respiratory issues, allergic reactions, and chronic skin conditions [14]. Inconsistent use of PPE, despite high awareness, suggests a difference between knowledge and practice. The reasons could include a lack of constant PPE supply, poor fit or comfort, or time pressures during emergencies. The effect is that PPE provision must go hand-in-hand with behavior-focused training, supportive supervision, and departmental accountability systems.

Exposure to biological hazards, particularly through needlestick injuries, remains a persistent issue. While similar trends are widely reported in the literature [14], the relatively lower incidence of reported infections in this study could be due to post-exposure prophylaxis availability, work overload, inadequate staffing during high-risk procedures, and delayed hazard reporting due to fear of stigma or reprisal. Strengthening infection prevention through regular drills, anonymous reporting systems, and routine access to post-exposure prophylaxis can mitigate these risks and protect both staff and patients.

Although psychological hazards were less frequently acknowledged, indicators of burnout suggest a silent yet impactful threat. Unlike studies from high-income countries that report high levels of workplace-related stress among healthcare workers [15], some underreporting here may reflect cultural stigmas surrounding mental health, limited awareness, or normalization of stress. The implication may be clear without proactive mental health interventions and open dialogue around psychological safety, the workforce remains vulnerable to emotional exhaustion and reduced quality of care. Institutional policies should include routine stress screening, mental health counseling, and wellness initiatives.

Another important insight is the generally high awareness of OHS policies and emergency procedures. This emphasizes the positive effects of targeted training on knowledge and self-efficacy in handling occupational risks [16]. However, having knowledge alone did not lead to safe practices. This was seen in the inconsistent use of PPE and delays in reporting hazards. This gap between what people know and what they actually practice is common in many healthcare systems in low- and middle-income countries [17]. It is often caused by challenges like poor supervision and a lack of follow-up. To improve this, occupational health and safety programs should include regular mentorship, refresher training, and systems that encourage team members to hold each other accountable.

In terms of knowledge, attitudes, and practices (KAP) surrounding OHS, the study found high overall awareness of safety protocols and emergency procedures, confirming prior research that links OHS training to improved safety culture [18]. Workers were generally confident in responding to emergencies and managing chemical exposures. However, the fact that safety discussions typically occurred only post-incident suggests a reactive rather than proactive safety culture. This reactive approach limits the potential for early hazard identification and prevention. To transform workplace safety, healthcare institutions must foster a culture of continuous dialogue, near-miss reporting, and preventive learning.

Importantly, variations in OHS awareness by job role and department were observed, with midwives and obstetric staff demonstrating higher knowledge than their peers. This supports earlier findings suggesting that professionals in high-risk and high-volume departments often receive more intensive and recurrent training [19]. On the other hand, the lower awareness among public health officers suggests a potential neglect of non-clinical units in safety programming. This implies that OHS training must be customized to departmental risks and job roles, ensuring equitable exposure to safety education across all staff categories.

The study also highlights the positive influence of safety training attendance on OHS knowledge and practices. This finding is consistent with international evidence that shows structured safety training improves hazard recognition, risk perception, and protective behavior [20]. Institutions must therefore prioritize regular, mandatory, and participatory training programs tailored to specific hazards and updated with emerging threats. Beyond skill development, such initiatives help embed a culture of collective responsibility and continuous improvement. This study highlights several areas requiring urgent attention, from strengthening PPE access and ergonomic design to enhancing mental health support and cultivating proactive safety cultures. The findings reinforce that improving OHS among healthcare workers is not a one-time event but a continuous process requiring policy commitment, tailored training, structural investment, and staff engagement. Future OHS programs must prioritize comprehensive, department-specific interventions that empower healthcare workers with both the knowledge and the means to protect themselves, ultimately leading to safer healthcare environments and better patient outcomes.

## 5. Conclusion

This study examines the occupational health and safety conditions of the Ashanti Regional Hospital, Ghana, by determining the common hazard groups, professionals’ knowledge, attitudes, and practices, as well as the determinant factors. With safety training and ergonomic intervention related to safe handling being the most needed, the results showed that biological hazards are most common, followed by physical, psychological, and chemical hazards. Although HCWs demonstrate high awareness of OHS and emergency preparedness protocols, compliance remains suboptimal, and an overall reactive stance towards safety needs to be addressed through proactive measures. OHS knowledge varies across departments and highlights the need for specific safety programs to target risks in specific healthcare units. This underscores that distributed safety training within the scope of occupational health and safety (OHS) will lead to a considerable increase in OHS discipline, reflecting that there must be a constant effort to keep a strong safety culture. These behaviours must be institutionally reinforced with proactive hazard prevention efforts, enforcement of this policy, and improved access to personal protective equipment (PPE), which is paramount in safeguarding healthcare workers and ensuring optimal patient care. Further research will be required to evaluate the long-term impacts of OHS training and interventions to enhance workplace safety.

## Data Availability

The data will be made available in a public repository upon publication.

## 6. Acknowledgements

We acknowledge all the study participants for their involvement in the study.

